# Digital otoscopy videos versus composite images: A reader study to compare the accuracy of ENT physicians

**DOI:** 10.1101/2020.08.17.20176131

**Authors:** Hamidullah Binol, M. Khalid Khan Niazi, Garth Essig, Jay Shah, Jameson K. Mattingly, Michael S. Harris, Charles Elmaraghy, Theodoros Teknos, Nazhat Taj-Schaal, Lianbo Yu, Metin N. Gurcan, Aaron C. Moberly

**Affiliations:** Center for Biomedical Informatics, Wake Forest School of Medicine, Winston-Salem, NC, USA; Department of Otolaryngology, Ohio State University, OH, USA; Case Western Reserve University School of Medicine, OH, USA; Otolaryngology and Comm. Sciences - Froedtert Hospital, WI, USA; University Hospitals Seidman Cancer Center, OH, USA; Department of Internal Medicine, Ohio State University College of Medicine, OH, USA; Department of Biomedical Informatics, Ohio State University, OH, USA

**Author notes:** **Corresponding author:** Hamidullah Binol **Postal address:** 486 N. Patterson Avenue, Winston-Salem, NC 27101, USA **Email address:** **ORCID:** https://orcid.org/0000-0003-2537-8280. **Funding**: The project described was supported in part by Award R21 DC016972 (PIs: Gurcan, Moberly) from National Institute on Deafness and Other Communication Disorders. The content is solely the responsibility of the authors and does not necessarily represent the official views of the National Institute on Deafness and Other Communication Disorders or the National Institutes of Health.

**Keywords:** Computer-assisted Diagnosis, Eardrum, Image stitching, Otoscope, Telemedicine

## Abstract

**Objectives:** With the increasing emphasis on developing effective telemedicine approaches in Otolaryngology, this study explored whether a single composite image stitched from a digital otoscopy video provides acceptable diagnostic information to make an accurate diagnosis, as compared with that provided by the full video.

**Methods:** Five Ear, Nose, and Throat (ENT) physicians reviewed the same set of 78 digital otoscope eardrum videos from four eardrum conditions: normal, effusion, retraction, and tympanosclerosis, along with the composite images generated by a *SelectStitch* method that selectively uses video frames with computer-assisted selection, as well as a *Stitch* method that incorporates all the video frames. Participants provided a diagnosis for each item along with a rating of diagnostic confidence. Diagnostic accuracy for each pathology of *SelectStitch* was compared with accuracy when reviewing the entire video clip and when reviewing the *Stitch* image.

**Results:** There were no significant differences in diagnostic accuracy for physicians reviewing *SelectStitch* images and full video clips, but both provided better diagnostic accuracy than *Stitch* images. The inter-reader agreement was moderate.

**Conclusion:** Equal to using full video clips, composite images of eardrums generated by *SelectStitch* provided sufficient information for ENTs to make the correct diagnoses for most pathologies. These findings suggest that use of a composite eardrum image may be sufficient for telemedicine approaches to ear diagnosis, eliminating the need for storage and transmission of large video files, along with future applications for improved documentation in electronic medical record systems, patient/family counseling, and clinical training.

**Level of Evidence:** Level 3

## Introduction

Clinical examination of the eardrum (tympanic membrane − TM) through handheld otoscopy is the most common diagnostic approach for TM pathologies^1^. Interpretation of the often brief glimpse of the TM obtained through the small viewing window requires extensive experience. With a growing need to develop effective telemedicine systems, which recently gained widespread attention during the COVID-19 pandemic^2^, novel methods to perform telemedicine otoscopy are needed. Previous studies have shown that telemedicine review of images is sufficiently accurate to use in burn^3–5^ and trauma care^6,7^. For otoscopy, one telemedicine approach is to apply digital otoscopy, during which a short video examination of the TM is recorded, which is reviewed by a telemedicine physician. Although there are no studies directly comparing otoscopic diagnoses based on a video clip as compared with a single digital image, our previous work led us to use videos^8^. There, diagnostic performance was compared between otoscopic single images and in-office microscopy. We included only images that were of sufficient focus/lighting, representing relatively ideal imaging conditions. Other authors have also noted insufficient image quality in a large percentage of their otoscopic still image databases, and/or the broad variability inherent across still images^9,10^.

The use of digital otoscopic video clips could overcome limitations imposed in real clinical settings: collecting a string of frames in a video could capture at least a few useful frames with sufficient focus and lighting, even in the setting of partially obstructing cerumen or a moving child. However, a major downside of video clips is that they require a large amount of storage space. A typical otoscopic video clip is 1440 × 1080 × 24 bits/pixels per frame, with between 200 and 1000 frames, contrasted with a single frame for a still image. Transferring large video clips, even after compression, can provide a major barrier to care in settings where internet bandwidth is limited^11–15^. Moreover, storage of videos within electronic medical records is not currently streamlined, and replaying videos for patients/families for counseling purposes is tedious.

Along similar lines, previous studies have been done to detect ear abnormalities by computer-based methods, requiring either manually extracting a single image from videos or capturing a single image with minimal glare/obstruction^9,10,16–19^. However, manually selecting a frame from a video is time-consuming and subject to high inter- and intra-reader variability^20–22^. A more sophisticated computerized method that creates a “composite” otoscopic image from a video should lead to a more useful final image, since a typical video comprises at least 200 frames, more than one of which may contribute information to the diagnostician.

We previously reported a computer-aided otoscopic frame selecting and stitching framework called *SelectStitch*^23^, in which a semantic segmentation-based framework automatically selects meaningful frames containing portions of the TM from videos, reducing irrelevant frames (e.g., those heavily blurred or having excessive cerumen). We then conducted a reader study with three Ear, Nose, and Throat (ENT) physicians who reviewed these composite *SelectStitch* images and compared them to composite images generated using the entire video (i.e., without frame selection, called *Stitch*) in terms of diagnostic decisions. Figure 1 provides an overview of *SelectStitch* and *Stitch*. We found that *SelectStitch* improved the diagnostic quality of composite images relative to *Stitch*. However, that study did not address several remaining questions important for the translation of this approach to the clinic and particularly to telemedicine settings, which are addressed in the current study:

**Figure 1.**
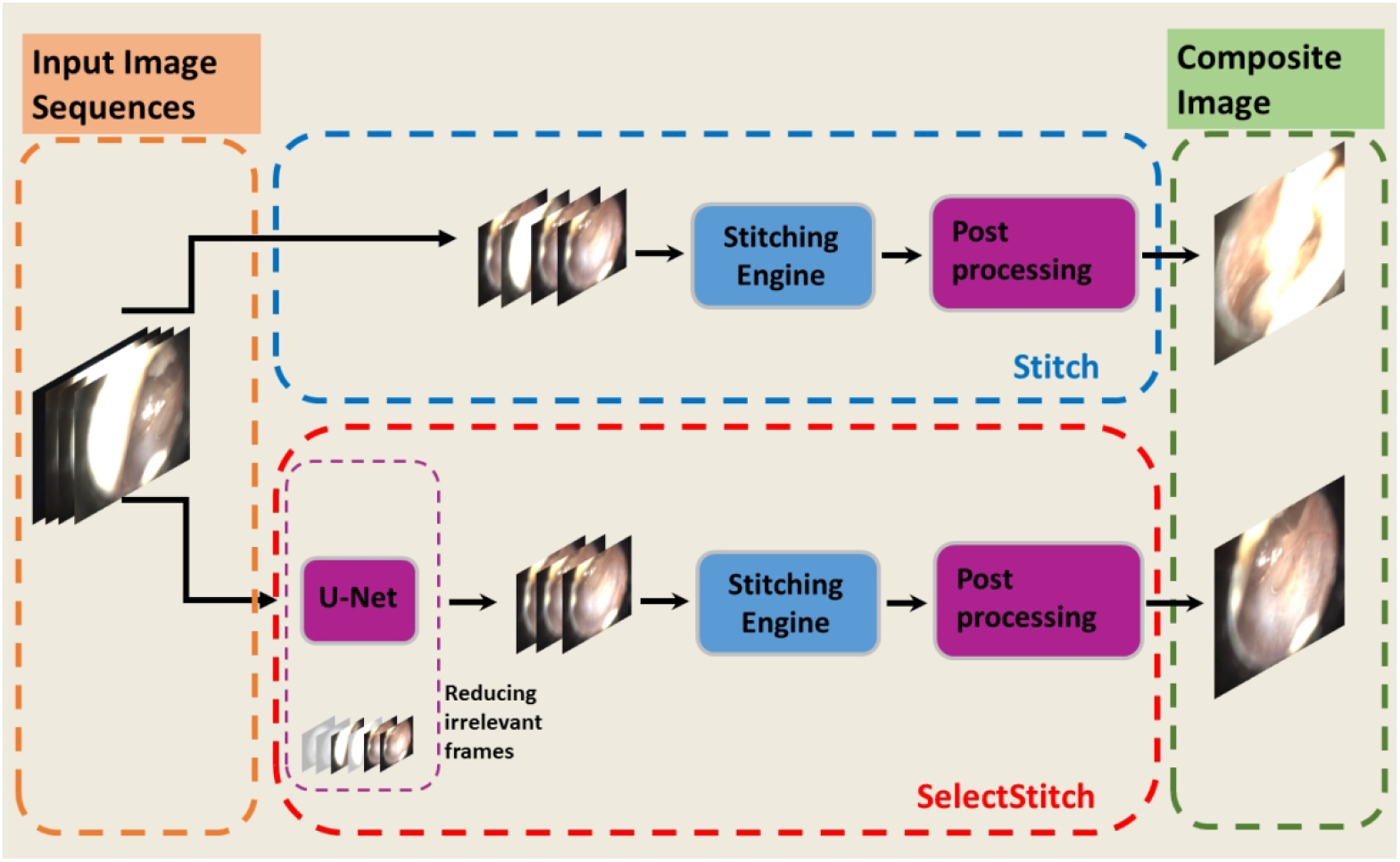
The process of *Stitch* and *SelectStitch*. In comparison to *Stitch, SelectStitch* possesses a deep learning based semantic segmentation step to reduce irrelevant frames from video sequences as described in^23^. These excluded frames include parts of the video with low quality (e.g., those heavily blurred or having an excessive amount of cerumen).

### (a) When reviewing SelectStitch composite images, what is the accuracy of diagnosis?

To answer this question, five ENTs reviewed 78 composite images and provided diagnoses, compared with a “true” diagnosis. For adult patients, the “true” diagnosis was based on digital otoscopy, supplemented with clinical microscopy as well as audiology testing (hearing testing and/or tympanometry). For pediatric patients, the “true” diagnosis was based on digital otoscopy, supplemented with microscopy in the operating room during placement of pressure equalization tubes. We also aimed to determine which pathologies were easiest and hardest to diagnose.

### (b) Is the accuracy of diagnosis for SelectStitch composite images different from Stitch images and from videos?

Because the *Stitch* technique generates composite images using all frames of a video, including redundant frames and frames of poor quality, we predicted that the diagnostic accuracy for *SelectStitch* images would be superior to the accuracy for *Stitch*. More importantly, we predicted that the diagnostic accuracy for *SelectStitch* would be equivalent to the accuracy for full video clips.

### (c) How does the level of confidence of ENTs for each diagnostic tool (SelectStitch, Stitch, and video) relate to diagnostic ability?

To answer this question, the five ENTs rated their level of confidence in making diagnoses for each type of pathology in each diagnostic tool condition.

### (d) What is the inter-reader variability of ENTs on diagnosing with the diagnostic tools?

As with any medical application, we expected that there would be inter-reader variability among ENTs, but that agreement would generally be relatively high.

## Materials and Methods

A database of high-resolution digital adult and pediatric videos, captured via a digital otoscope from ENT clinics and operating rooms, as well as in a primary care Medicine/Pediatrics setting, was created after Institutional Review Board (IRB) approval^8^. A high definition (HD) video otoscope (JEDMED Horus+ HD Video Otoscope, St. Louis, MO) was utilized^23^. The video frames were 1440 by 1080 pixels and were recorded in a MPEG 4 file format. In this study, 78 video clips from the database were used, selected if they only had one single diagnostic label associated. These videos consisted of 20 normal ears, 20 with middle ear effusions (serous or mucoid), 20 with TM retractions, and 18 with tympanosclerosis (i.e., myringosclerosis). Videos were excluded if they had low light throughout the video and/or if they did not contain a clear view of at least part of the TM. Where possible, we selected pediatric and adult videos as balanced (e.g., 10 pediatrics and 10 adults), except for tympanosclerosis for which there were 10 adult and eight pediatric videos.

An online diagnostic assessment tool was designed using SurveyMonkey, an online survey software. The video clips were hosted on Vimeo, and the composite images were uploaded to imgbox. An example of a question from our online survey is shown in Figure 2. Each sample (*Stitch* or *SelectStitch* composite image or video) was displayed on the screen, and the reader was asked to state the diagnosis (or normality). The order of presentation (video first or composite image first) was randomized to each clinician separated by four weeks (see Figure 3). If a reader viewed the video of a sample in the first survey, he/she read the *Stitch* and *SelectStitch* composite images (also in a randomized order) in the second survey. The cases from adult and pediatric patients were also mixed in each survey.

**Figure 2.**
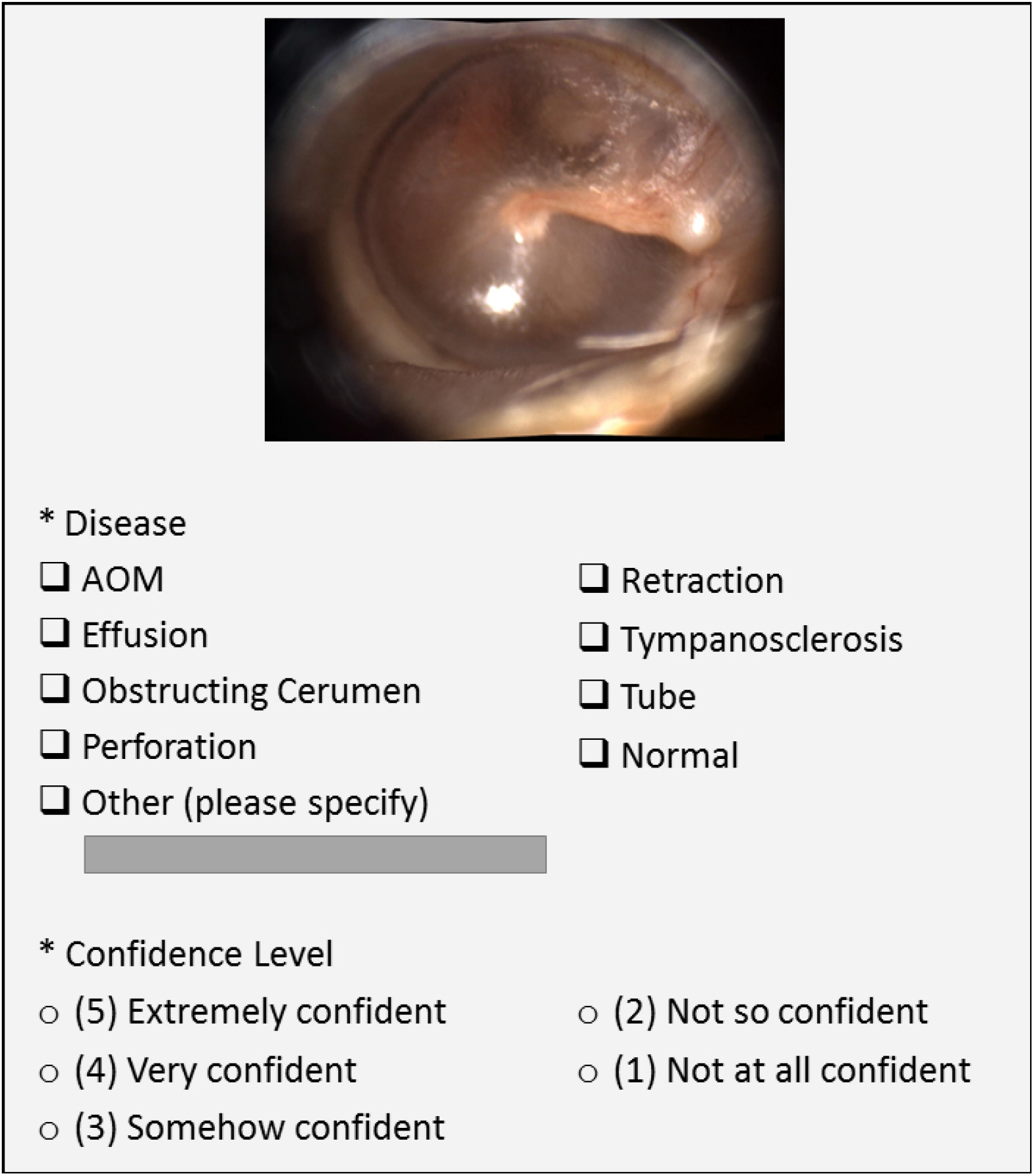
An example question from the online diagnostic survey. The readers are asked to make a diagnosis of the disease either from the video or a composite image (either produced by *Stitch* or *SelectStitch*). Readers can pick one or more of the choices. If their diagnosis is not included in any of the categories, they can pick the Other Category and enter their choice (e.g. monomeric TM). Readers are also asked their diagnostic confidence level using the Likert Scale with 5 being “extremely confident.”

**Figure 3.**
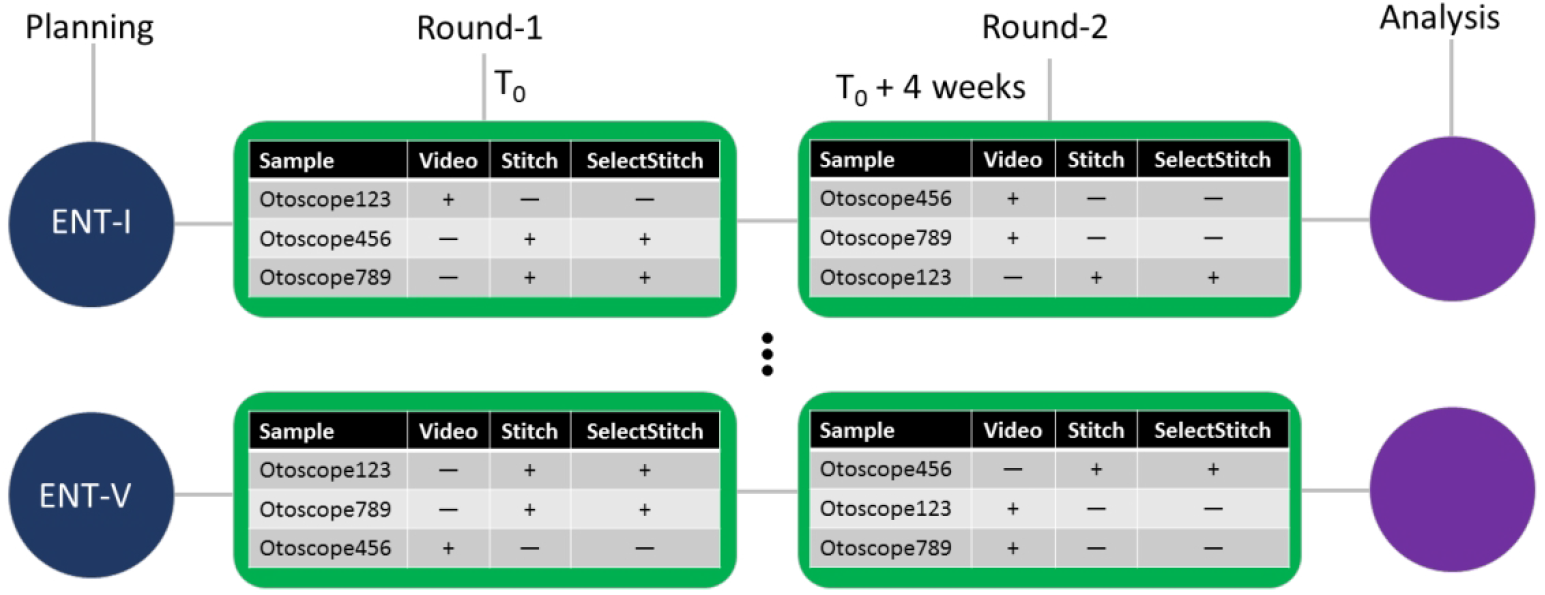
Summary of the rounds of the otoscope diagnosis survey for each reader (ENT-I through ENT-V). The order of the *Stitch* and *SelectStitch* composite images of the same sample were mixed in each survey. The cases from adult and pediatric patients were also mixed in each evaluation set.

At the completion of each survey, readers were asked to rate their degree of confidence in making each type of diagnosis, on a scale of 1 to 5, in which 1 indicated no confidence while 5 indicated extreme confidence. Five ENTs (authors ACM, GE, JS, JKM, and MSH; three neurotologists, one comprehensive otolaryngologist, and one pediatric otolaryngologist) were invited by email to complete the online assessment, and all completed the assessment after completing written informed consent.

### Statistical Analyses

Two different scoring strategies were applied to the survey answers. Although each sample had only one true diagnostic label, we did not restrict the readers regarding the number of diagnostic answers they could provide for each sample. Answers were scored using two different strategies. In Score-1, we scored the answers according to whether the reviewer provided the correct diagnostic answer as well as how many answers were given (e.g., the true label was effusion but the reviewer provided two diagnoses: effusion and tympanosclerosis). To compute accuracy using Score-1, an answer-weighting strategy was used: proportion 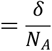 where N_A_ was the number of answers provided by the reviewer and δ was the binary output of answers, where the item received a 1 if any of the answers were correct and 0 otherwise. For example, if a reader selected two answers (N_A_ = 2) and one of them was correct (δ = 1), then the proportion (in percentage) for that particular sample would be 50%. It should be noted that this is not “accuracy” in a traditional sense. In contrast, for Score-2, an answer was accepted as correct if any diagnosis in the response matched the true label. Score-2 was computed as the percent of responses that contained a correct diagnosis, which is a relatively lenient approach to accuracy.

To study differences in scoring among diagnostic tools (video clips, *Stitch, SelectStitch*) and among the five ENT doctors, ordinal logistic regression was applied to both Score-1 and Score-2. Similar analysis was performed for studying the association between confidence level in scoring and scores. Wald tests were performed for comparison between diagnostic tools. Bonferroni method was used for multiple comparisons (e.g., α = 0.016 when adjusting for three comparisons). Kendall’s concordance was calculated to assess inter-reader agreement for each diagnostic tool, where a concordance of 0 suggests no inter-reader agreement and a concordance of 1 suggests perfect agreement.

## Results

### Question (a) What was the accuracy of diagnosis for SelectStitch?

As shown in Table 1, the overall proportions of diagnostic accuracy for *SelectStitch* images among ENTs varied between 46 and 62% for Score-1. The easiest and hardest categories to diagnose, respectively, were Tympanosclerosis (mean ± std: 69% ± 9) and Retraction (mean ± std: 39% ± 7).

**Table 1.** Proportion of correct diagnosis in percentages for each diagnostic category for each ENT physician (I through V) using Score-1 (S1) and Score-2 (S2) (%) using *SelectStitch*.

| Diagnostic Categories | ENT-I |  | ENT-II |  | ENT-III |  | ENT-IV |  | ENT-V |  | mean (SD) |  |
| --- | --- | --- | --- | --- | --- | --- | --- | --- | --- | --- | --- | --- |
|  | S1 | S2 | S1 | S2 | S1 | S2 | S1 | S2 | S1 | S2 | S1 | S2 |
| Normal | 55 | 55 | 55 | 55 | 65 | 65 | 85 | 95 | 55 | 55 | 63 (13) | 65 (17) |
| Effusion | 78 | 90 | 52 | 90 | 64 | 75 | 16 | 25 | 25 | 35 | 47 (26) | 63 (31) |
| Retraction | 38 | 55 | 45 | 65 | 29 | 40 | 48 | 70 | 37 | 50 | 39 (7) | 56 (12) |
| Tympanosclerosis | 78 | 94 | 57 | 78 | 67 | 78 | 78 | 83 | 65 | 89 | 69 (9) | 84 (7) |
| Average ( mean (SD) ) | 62 (19) | 74 (21) | 52 (5) | 72 (15) | 56 (18) | 65 (17) | 57 (32) | 68 (31) | 46 (18) | 57 (23) |  |  |

For Score-2, also shown in Table 1, the average accuracy rates of ENT doctors for *SelectStitch* suggested that the easiest category to diagnose was again Tympanosclerosis (mean ± std: 84% ± 7) and the hardest to diagnose was Retraction (mean ± std: 56% ± 12). Overall Score-2 accuracies among ENTs varied between 57 and 74%.

### Question (b) Did the accuracy of diagnosis differ among SelectStitch, Stitch, and video clips?

Similar tables of diagnostic accuracy are shown in the Appendix for *Stitch* (Appendix Table A) and video clips (Appendix Table B). For Score-1, overall, there was a significant difference in score among the three diagnostic tools at p value < 0.0001 (F value = 44.42). For paired comparisons, there was no significant difference between the video method and *SelectStitch* at p value = 0.9736; there was a significant difference between video method and *Stitch* (*Stitch* method scored less) at p value < 0.0001; and there was a significant difference between *SelectStitch* and *Stitch* (*Stitch* scored less) at p value < 0.0001 (Table 2). For Score-2, overall, there was also a significant difference in score among the three diagnostic tools at p value < 0.0001 (F value = 51.06). For paired comparisons, there was no significant difference between diagnostic accuracy for video clips and *SelectStitch* at p value = 0.9391; there was a significant difference between video and *Stitch* (*Stitch* scored less) at p value < 0.0001; and there was a significant difference between *SelectStitch* and the *Stitch* (*Stitch* scored less) at p value < 0.0001 (see Table 2). In summary, for both Score-1 and Score-2, diagnostic accuracy was equivalent for *SelectStitch* composite images and video clips, both of which were better than for *Stitch* images.

**Table 2.**
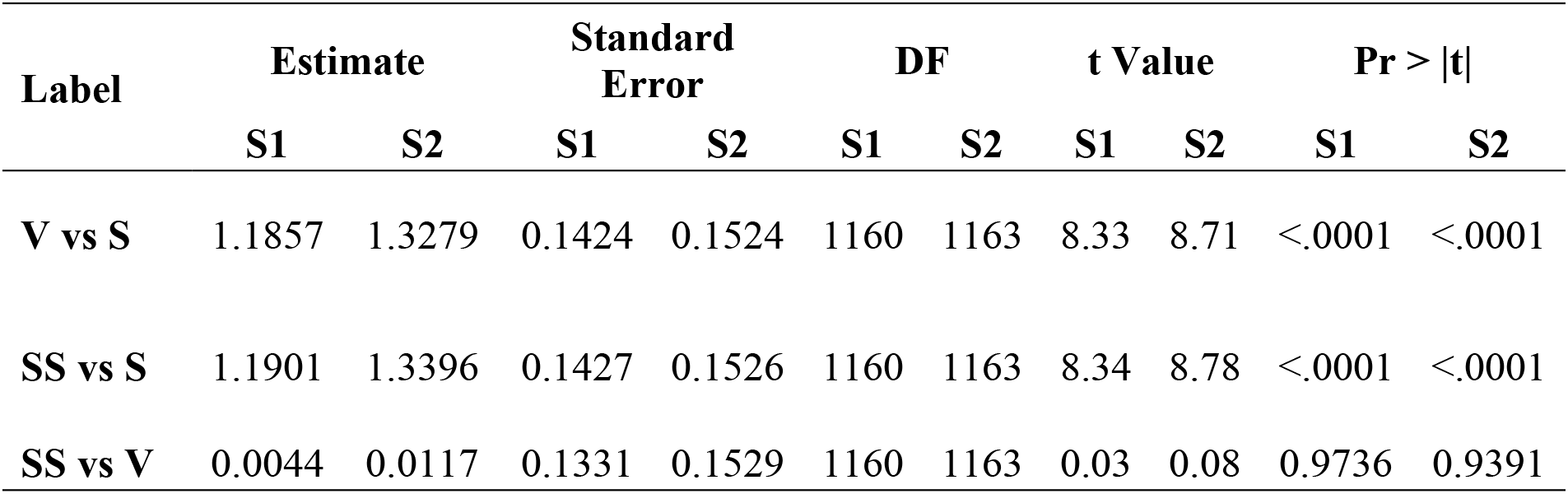
Comparisons of accuracy for the different diagnostic tools for Score-1 (S1) and Score-2 (S2). V = Video; S = *Stitch*; SS = *SelectStitch*.

| Label | Estimate |  | Standard Error |  | DF |  | t Value |  | Pr > \|t\| |  |
| --- | --- | --- | --- | --- | --- | --- | --- | --- | --- | --- |
|  | S1 | S2 | S1 | S2 | S1 | S2 | S1 | S2 | S1 | S2 |
| V vs S | 1.1857 | 1.3279 | 0.1424 | 0.1524 | 1160 | 1163 | 8.33 | 8.71 | <.0001 | <.0001 |
| SS vs S | 1.1901 | 1.3396 | 0.1427 | 0.1526 | 1160 | 1163 | 8.34 | 8.78 | <.0001 | <.0001 |
| SS vs V | 0.0044 | 0.0117 | 0.1331 | 0.1529 | 1160 | 1163 | 0.03 | 0.08 | 0.9736 | 0.9391 |

### Question (c) How did level of confidence of ENTs for each diagnostic tool relate to their diagnostic ability?

Associations between confidence level and Score-1 were examined for each diagnostic tool. For Stitch, this association was not significant (t value = −2.4) after Bonferroni correction (p value = 0.0168; α used for Bonferroni correction = 0.0167). For video clips, this association was positive and significant (t value = 3 and p value = 0.0027). Finally, for *SelectStitch*, this association was positive and significant (t value = 2.87 and p value = 0.0041). Next, we compared the magnitude of association between confidence and Score-1 among the three diagnostic tools. Results demonstrated no significant difference in this association between video clips and *SelectStitch* (p value = 0.9944). In contrast, the association between confidence level and Score-1 for video clips was significantly higher than that for *Stitch* (p value < 0.0001), and the association for *SelectStitch* was significantly higher than that for *Stitch* (p value < 0.0001). Similar findings were demonstrated for the associations between confidence level and Score-2 of each diagnostic tool. Specifically, for *Stitch*, the association was non-significant (t value = −2.3) after Bonferroni correction (p value = 0.0214; α used for Bonferroni correction = 0.0167). For video clips, the association was positive and significant (t value = 3.33 and p value = 0.0009). Finally, for *SelectStitch*, the association was positive and significant (t value = 3.19 and p value = 0.0015). Again, the magnitude of associations of confidence level and Score-2 were compared among the three diagnostic tools. There was no significant difference between video clips and *SelectStitch* (p value = 0.9936). However, again, the association was significantly higher for video clips than for *Stitch* (p value < 0.0001), and the association was significantly higher for *SelectStitch* than for *Stitch* (p value < 0.0001). In summary, the associations between diagnostic accuracy (using both Score-1 and Score-2) and confidence level were significant only for *SelectStitch* and video clips, and these associations were of similar magnitude.

### Question (d) What was the inter-reader variability of ENTs with each diagnostic tool?

Kendall’s concordance was used for assessing inter-reader agreement. For Score-1, concordance was 0.5778 for *Stitch*, 0.4096 for video clips, and 0.4779 for *SelectStitch*. For Score-2, concordance was 0.584 for *Stitch*, 0.4312 for video clips, and 0.3529 for *SelectStitch*. These Kendall’s concordance values are all moderate in magnitude.

## Discussion

Telemedicine approaches have recently been highlighted during the COVID-19 pandemic. Even before then, telemedicine started to gain increasing attention in Otolaryngology^24–26^. Otoscopy is well-suited to the telemedicine approach^27,28^, as long as a sufficient image of the TM can be obtained. One way to optimize a sufficient image is to collect a short video clip of the examination. However, this results in a digital file that is relatively large, posing a barrier to both storage and transfer, especially in remote settings^27,29^. We hypothesized that computer-assisted creation of a composite image would maintain equivalent diagnostic utility and physician confidence during diagnosis.

Results demonstrated that the accuracies of ENTs in making diagnoses from *SelectStitch* images were equivalent to those made when reviewing the full videos, regardless of how diagnostic accuracy was determined (the stringent Score-1 vs the lenient Score-2). The overall average accuracies of ENTs (specifically for Score-2) were from 57 to 74%, a range that is similar to our previous study^8^. However, diagnostic accuracy depended largely on the type of pathology. For example, experts were 84% accurate in diagnosing tympanosclerosis, which has some distinguishing features (i.e., discrete areas of white plaque). In contrast, accuracy was lowest for the diagnosis of TM retraction, which can be a fairly subtle finding. Nonetheless, the most important finding of this study was that there were no significant differences in diagnostic accuracy between *SelectStitch* composite images and the full video clips. In contrast, *Stitch* composite images, which were constructed using all available frames of a given video, led to much poorer diagnostic accuracy than either *SelectStitch* or full video clips. This is a highly significant finding, because it suggests that single *SelectStitch* images provide details that are of equal diagnostic value to full video clips for expert reviewers.

Additionally, diagnostic accuracy and diagnostic confidence level were associated for both the *SelectStitch* and full video clips, while no association was found for *Stitch* images, which is interesting in light of previous work that demonstrated an overall weak relationship between diagnostic accuracy and confidence in ear experts^8^. This finding is important, because treatment decisions are often impacted by level of diagnostic confidence of the clinician. For example, a physician may need to feel confident of providing a diagnosis of “normal” in order to choose not to prescribe antibiotics for a patient presenting with otalgia. Moreover, inter-reader agreement in this study was generally only moderate in magnitude, providing further motivation for the need to develop methods to improve the objectivity of making ear diagnoses^16,17^.

This study has several limitations. First, only a subset of pathologies was included in the survey, while several important ear pathologies were excluded, such as acute otitis media. This was a result of small numbers of videos of some pathologies in our current database. Also, only videos of relatively high quality/lighting were included. Another limitation is that within-reader agreement was not evaluated. Lastly, each reader used his/her own computer monitor to evaluate the images and videos. Those monitors were likely of different makes, models, and resolutions. All of these factors could contribute to differences in diagnostic abilities. On the other hand, our approach was likely ecologically valid; in various telemedicine settings, a variety of monitors will be used.

Although the emphasis of this study was to provide support for the value of *SelectStitch* composite images in potential telemedicine settings, there are other scenarios for which an otoscopic composite image is likely preferable over a video clip. For example, current electronic medical record systems are more amenable to inclusion of photo-documentation in patient charts, as compared with video examinations. Additionally, the ability to show a patient or parent a simple composite image of a TM would improve counseling, such as in providing visual confirmation of a normal ear in a child with otalgia, which may help decrease over-prescription of oral antibiotics.

## Conclusion

Results of this study demonstrated that computer-aided *SelectStitch* composite images provide equivalent visual information as digital otoscopic video clips for ear experts to make diagnoses of different types of pathologies. Diagnostic accuracy was also found to be associated with diagnostic confidence level, and inter-reader agreement was moderate. Future studies will be required to evaluate a more diverse set of ear pathologies, as well as using videos collected under less ideal focus and lighting conditions.

## Data Availability

The data that support the findings of this study are available from the corresponding author, HB, upon reasonable request.

## Acknowledgments

The authors would like to thank Emily Luo and Benjamin Liu for curating videos and online surveys for this study.

## Appendix

**Appendix Table A.** Proportion of correct diagnosis in percentages for each diagnostic category for each ENT physician (I through V) using Score-1 (S1) and Score-2 (S2) (%) using *Stitch*.

| Diagnostic Categories | ENT-I |  | ENT-II |  | ENT-III |  | ENT-IV |  | ENT-V |  | mean (SD) |  |
| --- | --- | --- | --- | --- | --- | --- | --- | --- | --- | --- | --- | --- |
|  | S1 | S2 | S1 | S2 | S1 | S2 | S1 | S2 | S1 | S2 | S1 | S2 |
| Normal | 15 | 15 | 5 | 5 | 10 | 10 | 33 | 35 | 20 | 20 | 17 (11) | 17 (11) |
| Effusion | 68 | 70 | 31 | 40 | 55 | 55 | 10 | 10 | 13 | 15 | 35 (26) | 38 (26) |
| Retraction | 39 | 45 | 38 | 45 | 23 | 25 | 36 | 40 | 20 | 35 | 31 (9) | 38 (8) |
| Tympanosclerosis | 64 | 67 | 28 | 33 | 38 | 39 | 49 | 50 | 43 | 50 | 44 (13) | 48 (13) |
| Average ( mean (SD) ) | 46 (24) | 49 (25) | 25 (14) | 31 (18) | 31 (19) | 32 (19) | 31 (16) | 33 (17) | 23 (13) | 30 (16) |  |  |

**Appendix Table B.** Proportion of correct diagnosis in percentages for each diagnostic category for each ENT physician (I through V) using Score-1 (S1) and Score-2 (S2) (%) using video clips.

| Diagnostic Categories | ENT-I |  | ENT-II |  | ENT-III |  | ENT-IV |  | ENT-V |  | mean (SD) |  |
| --- | --- | --- | --- | --- | --- | --- | --- | --- | --- | --- | --- | --- |
|  | S1 | S2 | S1 | S2 | S1 | S2 | S1 | S2 | S1 | S2 | S1 | S2 |
| Normal | 60 | 60 | 55 | 55 | 70 | 70 | 93 | 95 | 29 | 30 | 61 (23) | 62 (24) |
| Effusion | 84 | 90 | 68 | 85 | 70 | 75 | 23 | 25 | 16 | 30 | 52 (31) | 61 (31) |
| Retraction | 48 | 55 | 53 | 70 | 35 | 45 | 59 | 60 | 49 | 70 | 49 (9) | 60 (11) |
| Tympanosclerosis | 83 | 89 | 65 | 78 | 72 | 48 | 80 | 83 | 76 | 94 | 75 (7) | 78 (18) |
| Average ( mean (SD) ) | 68 (18) | 73 (19) | 60 (7) | 72 (13) | 62 (18) | 67 (15) | 64 (31) | 65 (31) | 42 (26) | 55 (31) |  |  |

